# m6A RNA methylation regulators contribute to progression and impact the prognosis of breast cancer

**DOI:** 10.1101/2020.10.13.20212332

**Authors:** Wenjie Jiang, Minglong Dong, Zebin Hu, Kaidi Wan, Han Wang

**Affiliations:** School of Information Science and Technology, Northeast Normal University, Changchun 130117, China; Institute of Computational Biology, Northeast Normal University, Changchun 130117, China; Department of Computer Science, College of Humanities & Sciences of Northeast Normal University, Changchun 130117, China

**Keywords:** M6A RNA methylation, BRCA, LASSO, Prognostic

## Abstract

N6-methyladenosine (*m*^6^*A*) is the most commonly modified form of mRNA. *M*^6^*A* RNA methylation regulators are proved to be expressed clearly in some cancers by plenty of studies. Moreover, they also are proved to be indirectly involved in the growth of cancers. However, it remains unclear that the role of *m*^6^*A* RNA methylation regulator in the prognosis of breast cancer (BRCA). The data that we used in this study is the mRNA expression data obtained from the corresponding clinical information and the Tumor Genome Atlas (TCGA) database. And the goal we used the Wilcoxon rank-sum test was to evaluate the difference in the expression of *m*^6^*A* RNA methylation regulators in the normal group and the tumor group, and analyze the correlation between *m*^6^*A* RNA methylation regulators. We identified two subgroups of BRCA (cluster1 and 2) by using the K-mean algorithm and analyzing the correlation between clinic information and subgroups. The LASSO regression model then was used to figure out three *m*^6^*A* RNA methylation regulators, namely YTHDF3, ZC3H13, and HNRNPC. The riskScore of each patient was calculated according to the regression coefficients of the three *m*^6^*A* RNA methylation regulators. Base on the riskScore, we divided the patients into two groups, the high-risk group, and the low-risk group. After analyzing, we found that the overall survival rate (OS) of the low-risk group was higher than that of the other group. We conducted a univariate and multi-factor independent prognostic analysis of riskScore and three *m*^6^*A* RNA methylation regulators, and found that riskScore has a significant correlation with BRCA.

In conclusion, the *m*^6^*A* RNA methylation regulator is closely related to the development of BRCA, and the prognostic factor riskScore obtained from the regression of the expression of the three *m*^6^*A* RNA methylation regulators in the human body are likely to guide the individualization of BRCA patients A useful prognostic biomarker for treatment.

## Introduction

BRCA occurring in the epithelial tissue of the breast is a malignant tumor, and 99% of the patients are women and the rest are men ^(1)^, so it is considered to be the main cause of death in gynecological tumors. Due to advances in early screening and anti-cancer strategies, BRCA treatment^(2)^ has improved significantly. However, BRCA still shows a high recurrence rate^(3)^, and the cause is not yet clear. We must establish a model to deal with the prognosis of BRCA patients.

*m*^6^*A* (N6-methyladenosine, 6-methyl adenine) is the most common post-transcriptional modification of eukaryotic mRNA, accounting for 80% of RNA methylation modifications^(4)^. As early as the 1970s, people had discovered *m*^6^*A* modifications in the mRNA and lncRNA of eukaryotes^(5)^. It is known that in most eukaryotes, the methylation modification that occurs in the mRNA region of 5’UTR plays a significant role in mRNA splicing, editing, stability, degradation, and polyadenylation^(6-8)^; while the modification of the 3’UTR region Methylation modification contributes to the nuclear transport of mRNA, the initiation of translation, and the structural stability of the mRNA together with the polyA binding protein ^(9)^. *m*^6^*A* methylase is widely involved in mammalian development, immunity, tumor generation and metastasis, stem cell renewal, fat differentiation, and other life processes. METTL3 and METTL14 can promote the self-renewal of hematopoietic stem cells, and regulate *m*^6^*A* levels in the body in glioblastoma and liver cancer; FTO is the first *m*^6^*A* demethylase discovered, both single-stranded, double-stranded RNA and DNA It has the function of demethylation and mainly acts on the *m*^6^*A* site within the gene, as an oncogene to promote the occurrence of leukemia; besides, FTO is closely related to fat development; ALKBH5 can enhance the expression of its target gene FOXM1 and maintain the growth of glioma stem cells Growth; YTHDF1, YTHDF3, and YTHDC2 promote protein synthesis by improving the efficiency of *m*^6^*A* translators, while YTHDF2, YTHDF3, and YTHDC2 specifically recognize *m*^6^*A* mRNA and mediate mRNA degradation ^(10-14)^. More and more studies have proved that the development of cancer is deeply influenced by *m*^6^*A* RNA methylation regulators, but people have no further details about the relationship between *m*^6^*A* RNA methylation regulators and BRCA. Many studies construct prognostic signatures based on miRNAs ^(15)^ and long-chain non-coding RNAs (lncRNAs) ^(16)^ to predict the prognosis of BRCA. However, the prognostic signature that based on *m*^6^*A* RNA methylation modulators has not been constructed on the prediction of the prognosis of BRCA.

In this study, we analyzed the differences in the correlation among these *m*^6^*A* RNA methylation regulators and the expression of 15 *m*^6^*A* RNA methylation regulators^(17)^ in BRCA are based on the data which is collected from the Cancer Genome Atlas (TCGA) database ^(18)^. We constructed a regression mode with 3 *m*^6^*A* methylation regulators selected after Cox univariate analysis and LASSO regression analysis with p-value<0.1. Then, we studied the prognostic value of riskScore in BRCA, finding *m*^6^*A* RNA methylation regulators take a significant part in the progression of BRCA. Moreover, their riskScore can be used as the prognosis of BRCA patients.

## Materials and methods

### Data Acquisition

We used the data in the TCGA database (https://cancergenome.nih.gov/ to construct the RNA-seq transcriptome data of 1,222 BRCA patients and the corresponding clinicopathological and prognostic information.

### m^6^A RNA methylation regulators selection

In BRCA, there are 15 *m*^6^*A* -related genes and all of them have been identified as *m*^6^*A* methylation regulators. These 15 *m*^6^*A* -related genes have not been analyzed systematically in BRCA but have been analyzed systematically in pancreatic cancer. The BRCA mRNA expression data of 15 *m*^6^*A* -related genes were collected from the TCGA database and the expression of 15 *m*^6^*A* -related genes and clinical-pathological variables in BRCA patients were used to analyze the relationship among them.

### Bioinformatics analysis

Use the R “corrplot” package ^(19)^ to study the interactions between *m*^6^*A* RNA methylation regulators, and then use the k-mean clustering algorithm ^(20)^ to classify tumors for further analysis. In order to define the prognostic values of *m*^6^*A* RNA methylation regulators in BRCA patients, then we used the data in the TCGA database to perform Cox univariate analysis^(21)^ and selected *m*^6^*A* RNA methylation regulators with p-value<0.1 as the prognostic-related gene to construction model, and finally developed riskScore by LASSO regression algorithm^(22)^, which bases on a linear combination of regression coefficients and its expression level. The riskScore calculation method is as follows:

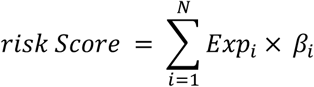

Based on the median value of riskScore, we divided these BRCA patients into high-risk groups and low-risk groups, and the survival curve of the patients was drawn, and then the correlation between riskScore and clinical information was analyzed. Finally, riskScore and other clinical information are used as prognostic factors for independent prognostic analysis, and p-value<0.05 indicates statistically significant correlation.

## Result

### The situation of *m*^6^*A* RNA Methylation Regulators in BRCA

First the expression levels of 15 *m*^6^*A* RNA methylation regulators in 1109 BRCA cases and 113 normal cases in the TCGA dataset were compared. Among them, the expressions of 11 *m*^6^*A* RNA methylation regulators (FTO, YTHDF1, HNRNPC, HNRNPA2B1, KIAA1429, ZCH13, YTHDC1, METTL14, WTAP, RBM15, and YTHDF2) are significantly related (**figure1A, B)**. We speculate it may be an inherent feature that the change in the ratio of *m*^6^*A* RNA methylation regulators, characterizing the differences between individuals. **Figure 1C** has shown the correlations between different *m*^6^*A* RNA methylation regulators. Among these 15 *m*^6^*A* RNA methylation regulators, lots of positive correlations, and negative correlations can be recognized.

**Figure1.**
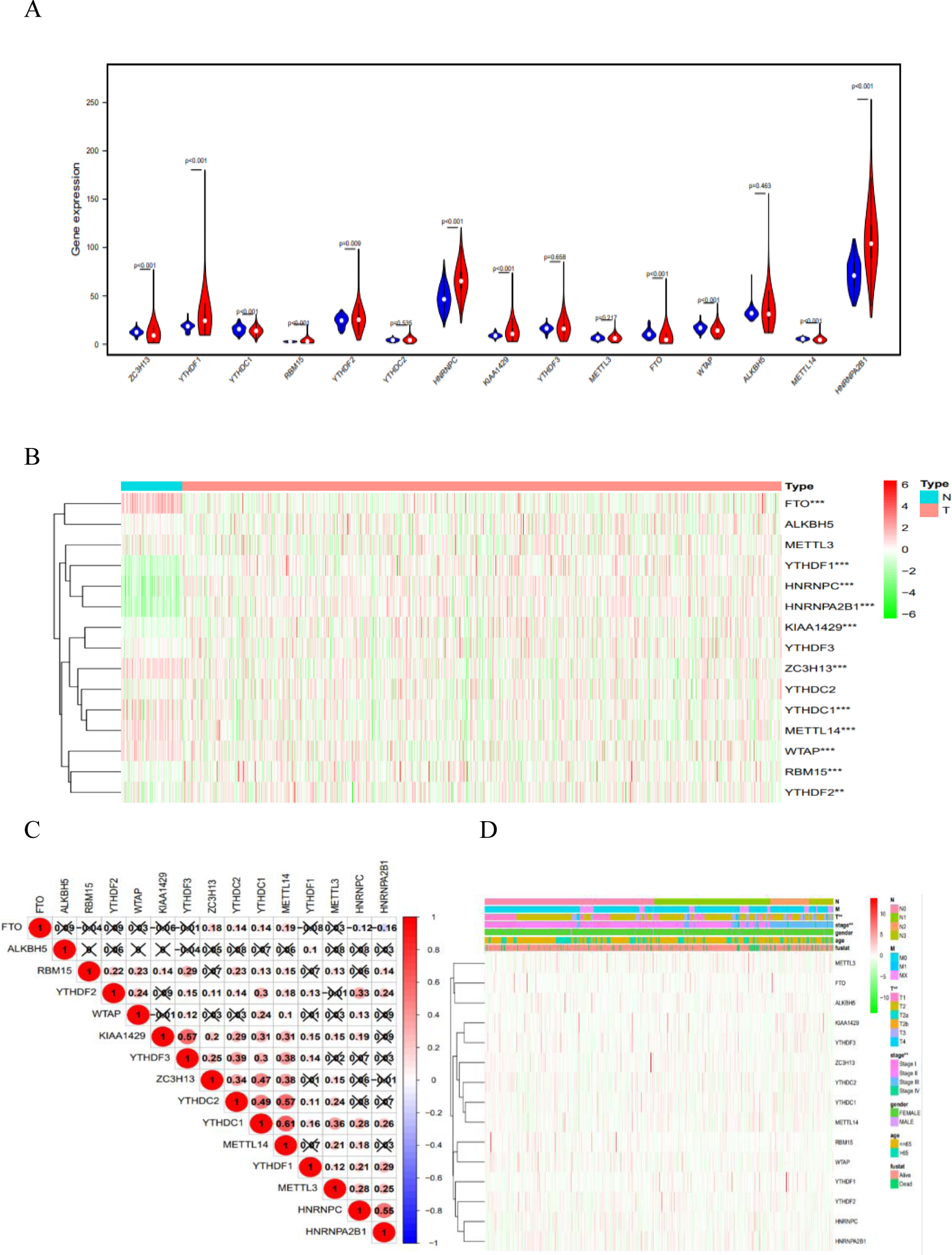
The expression levels and correlations analysis of *m*^6^*A* RNA Methylation Regulators in BRCA. (A) Visualize the expression levels difference of 15 genes in breast cancer patients (red) and normal people (blue). (B) The expression levels of 15 *m*^6^*A* RNA methylation regulators in BRCA. (C) Visualize the correlations among these 15 genes in BRCA. (D) Analyze the relationship between pathological characteristics and each *m*^6^*A* RNA methylation regulator

For example, the YTHDC1 gene and the YTHDF1 gene can be seen as a positive correlation as they have the strongest correlation, but the HNRNPA2B1 gene and the FTO gene would get a negative correlation. As shown in **Figure 1C**, The YTHDF1 gene is most probable to be up-regulated as the YTHDC1 gene is up-regulated, but the HNRNPA2B1 gene is most probable to be down-regulated when the FTO gene is up-regulated. The relationship between the pathological characteristics of BRCA, including age, gender, grade, status, stage, T status, M status, and N status and each *m*^6^*A* RNA methylation regulator was also systematically studied, finding that the *m*^6^*A* RNA methylation regulator indeed has a relationship with the pathological characteristics of BRCA (**Figure 1 d**).

### Two Clusters of BRCA Identified In Consensus Clustering of *m*^6^*A* RNA Methylation Regulators

We took 113 normal BRCA samples and used the R” ConsensusClusterPlus” package to group 1109 cancer tissues. The CDF value of k = 3 in the TCGA data set seems to be small from the perspective of the expression similarity of *m*^6^*A* RNA methylation regulators (**Figure 2B,C)**, but the correlation between the groups gets higher, and the number of samples gets smaller after being divided into three groups. Thus, the cancer tissues were grouped into two different kinds (**Figure 2A**).

**Figure2.**
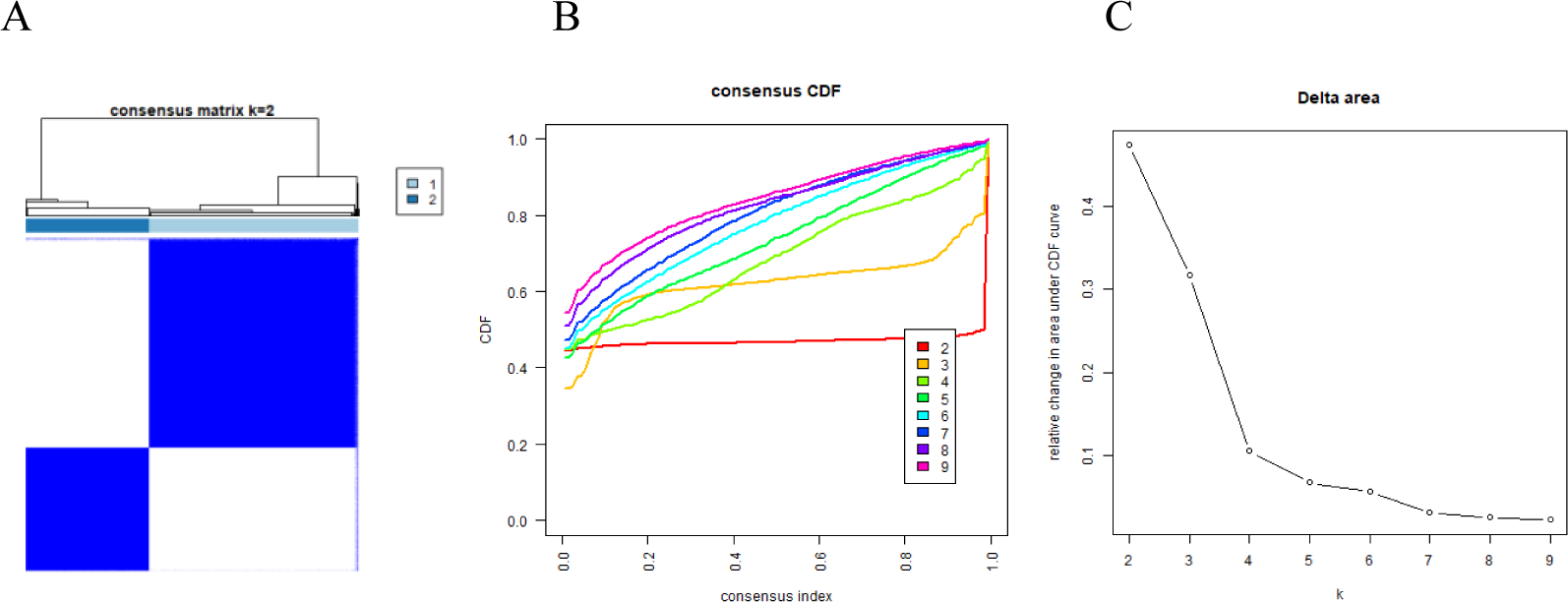
The analysis of clusters identification. (A) the matrix in Consensus clustering with k = 2; (B) cumulative distribution function (CDF) values in cluster when k equals 2 to 9; (C) the relative change in area under the CDF curve when k equals 2 to 9.

### The categories determined by consensus clustering are closely related to clinical outcomes and clinical pathology feature

The cluster results and OS curves of 1109 BRCA patients were analyzed to get better understanding of the cluster results and clinical efficacy and clinicopathological characteristics. In the cluster 2 subgroup, we discovered that most of the *m*^6^*A* RNA methylation regulators were highly expressed.

Moreover, compared the cluster 1 subgroup with the cluster 2 subgroup, we found that cluster 2 subgroup was apparently correlated with stage and T status at the time of diagnosis (**Figure 3)**.

**Figure3.**
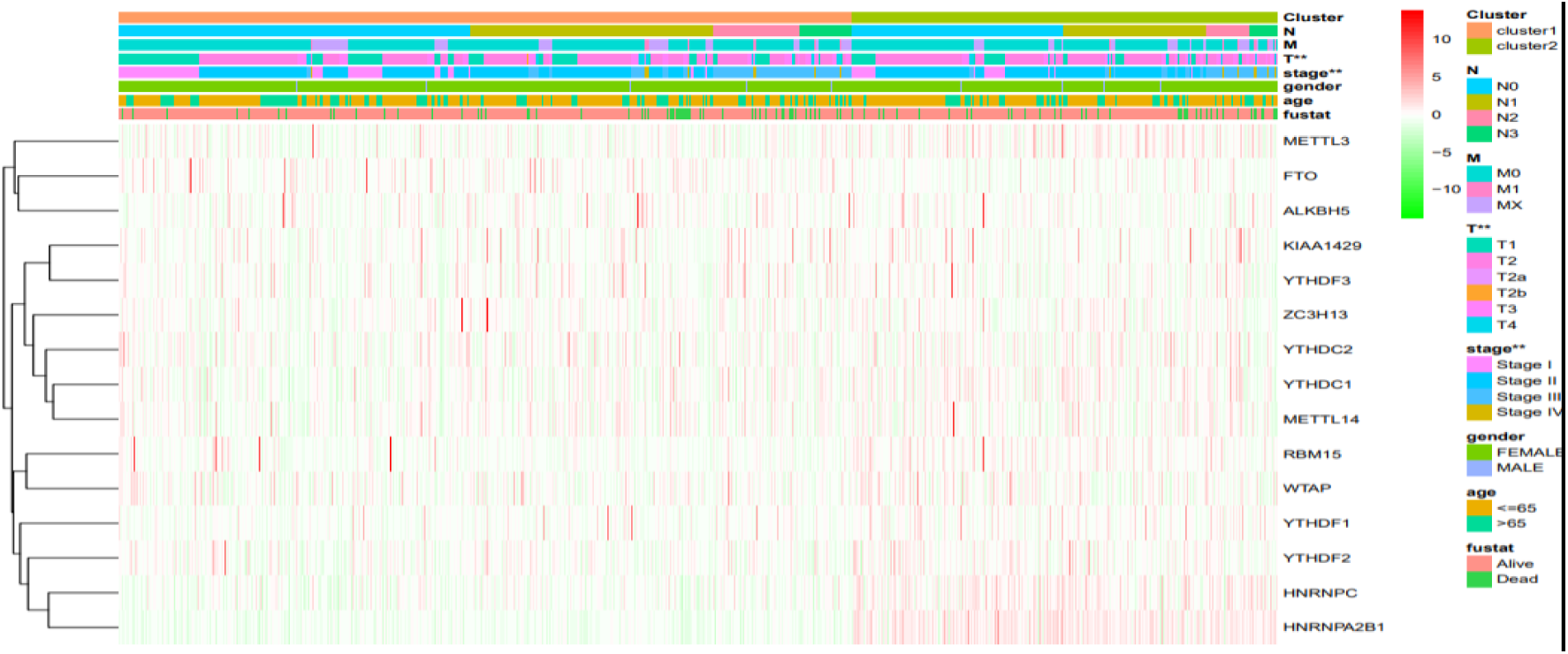
The heat map shows the expression of all m6A RNA methylation regulators and the distribution of clinicopathological variables between the high-risk and low-risk groups.

### prognostic value of *m*^6^*A* RNA methylation regulators and RiskScore

To get better understanding of the prognostic role of *m*^6^*A* RNA methylation modulator in BRCA, we performed univariate Cox regression analysis for expression levels of them. After analyzing, the results showed that the higher the expression of ZC3H13 and YTHDF3 is, the lower the survival rate of BRCA patients is, and the lower the expression of HNRNPC is, the higher the survival rate of patients is (**Figure 4A)**. For predicting the clinical outcome of *m*^6^*A* RNA methylation regulator on BRCA, the Least Absolute Contraction and Selection Operator (LASSO), and Cox regression algorithm was applied to 15 genes in the TCGA dataset. Based on the minimum criteria, we selected three genes (YTHDF3, ZC3H13, and HNRNPC) to construct riskScore and used the coefficients gained from the LASSO algorithm to calculate the riskScore of the TCGA dataset (**Figure 4B, C)**. Then BRCA patients in the TCGA data set were divided into high-risk and low-risk groups according to the median riskScore to explore the impact of three-gene risk characteristics on prognosis. The results showed that BRCA patients in the high-risk group had poorer survival (**Figure 4D)**.

**Figure4.**
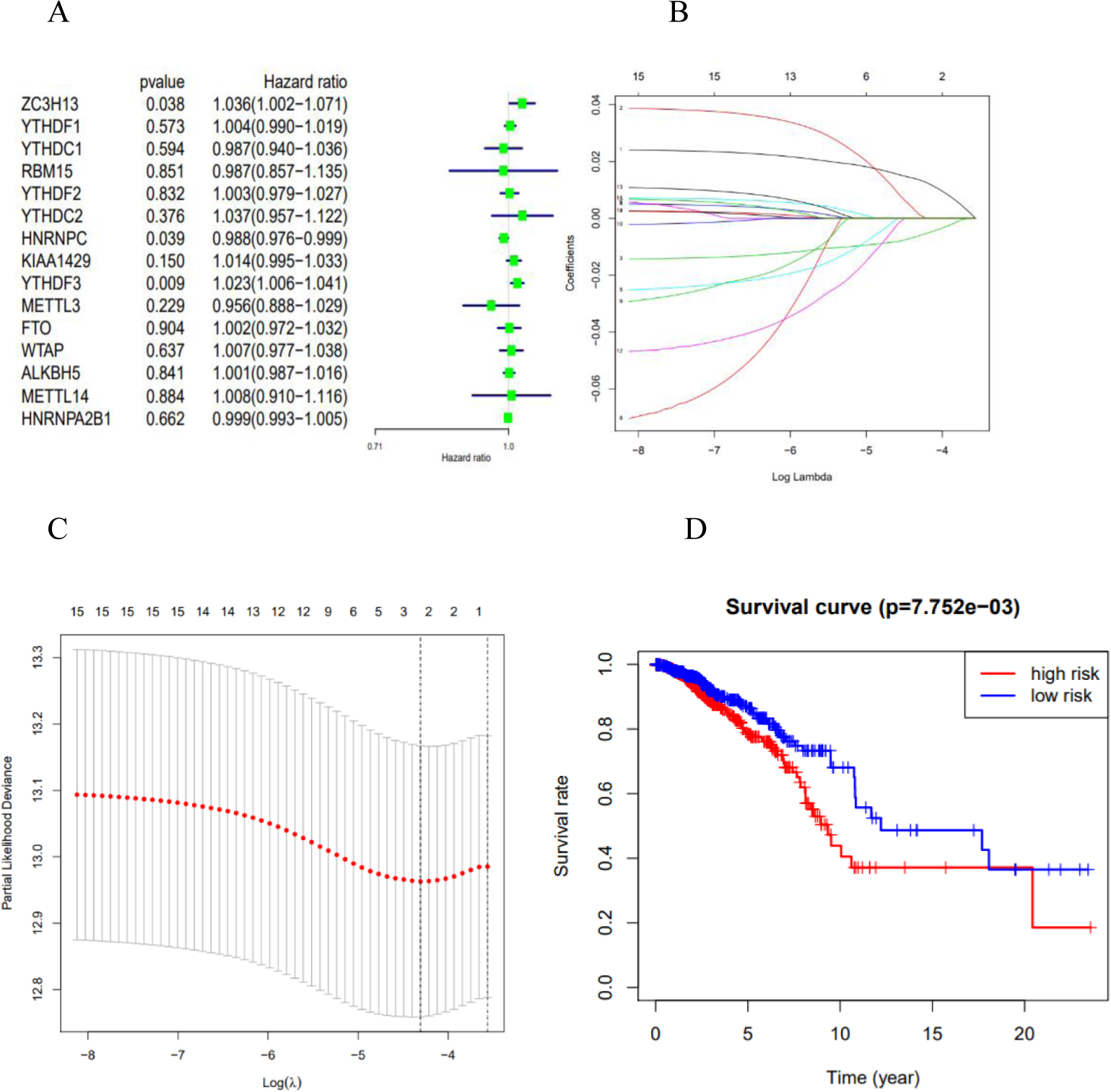
the impact on the prognostic of BRCA. (A) univariate Cox regression analysis for expression levels of m^6^A RNA methylation regulators; (B, C) the LASSO algorithm on three selected m^6^A RNA methylation regulators; (D) the survival curve in two clusters.

### Breast cancer prognostic risk score has a strong correlation with clinicopathological characteristics

The pathological characteristics including age, Stage status, T status, and N status and correlations of three *m*^6^*A* RNA methylation regulators in high-risk and low-risk groups were systematically studied to get better understanding of the clinical outcomes of BRCA in high-risk populations, finding that the selected three *m*^6^*A* RNA methylation regulators are related to the pathological characteristics of BRCA (**Figure 5A)**. Also, the expression of HNRNPC in the high-risk group patients was lower than these in the low-risk group, but the ratio of YTHDF3 and ZC3H13 was higher (**Figure 5A)**. To get better understanding of the relationship between riskScore and BRCA patients, we first made a ROC curve for predicting the riskScore and 3-year survival rate of BRCA patients. The results showed that the riskScore can predict the 3-year survival of BRCA patients Rate (AUC = 0.677) (**Figure 5B)**. In order to determine whether the risk characteristics are independent prognostic indicators, we then performed univariate and multivariate Cox regression analysis on the TCGA data set. The univariate and multivariate Cox regression analysis shows that the risk of patient death increase as the riskScore, age, stage status, T status, M status, and N status increase (**Figure 5C, D)**.

**Figure5.**
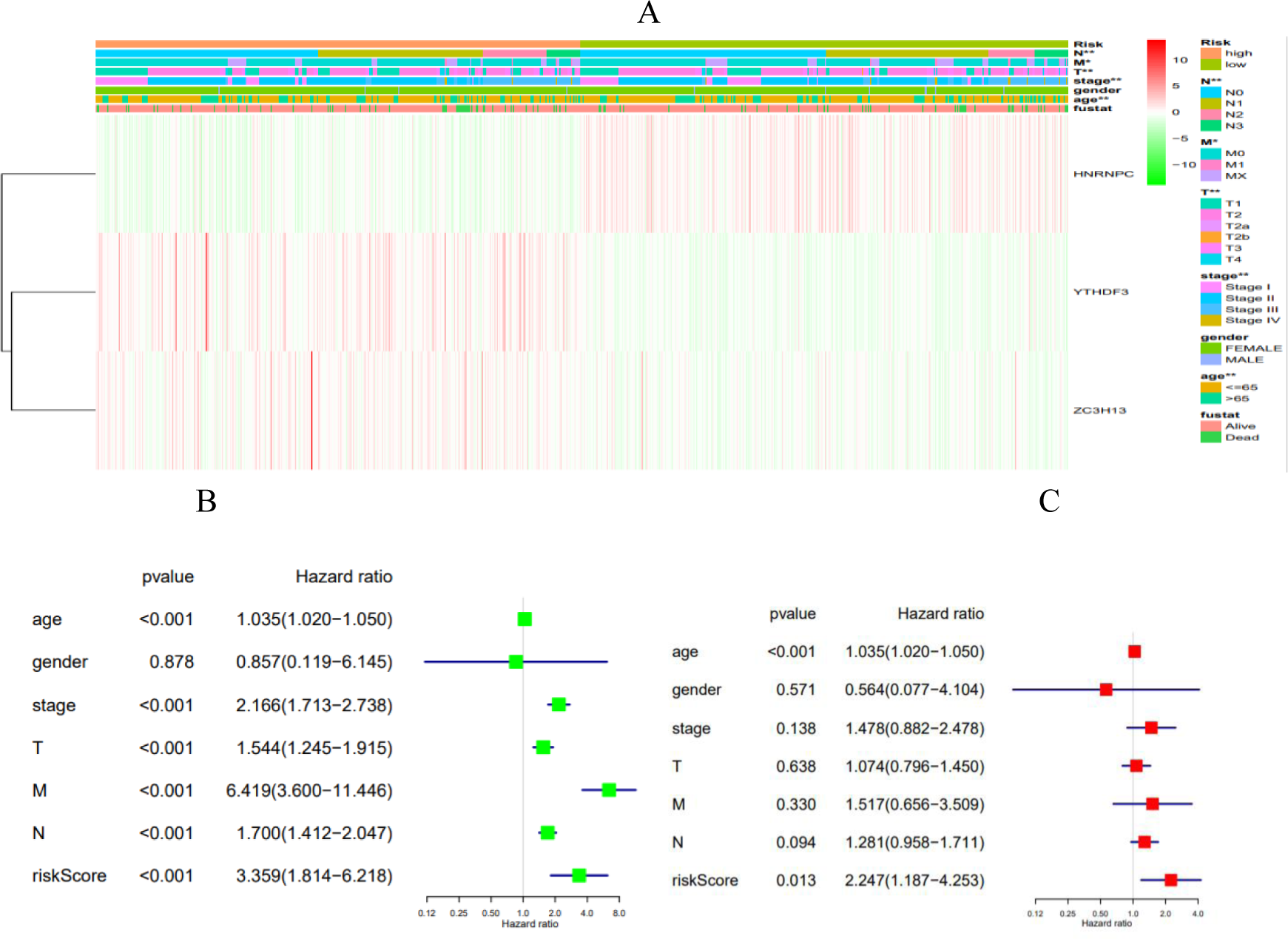
the result on three selected m^6^A RNA methylation regulators. (A) heat map that shows the expression levels of three selected regulators and the distribution of clinicopathological characteristics (B) univariate Cox regression analysis for clinicopathological characteristics with OS (C) multivariate Cox regression analysis for clinicopathological characteristics with OS.

## DISCUSSION

BRCA is one of the largest numbers of newly diagnosed cancer cases ^(23)^, and Women who die of BRCA are second only to lung cancer ^(24)^. Factors affecting the risk of BRCA development include history of breast disease in oneself and his family, age, Radiation exposure, race, and dietary factors. In 2016, the American Cancer Society predicted that approximately 249,260 new cases will be diagnosed, and 40,890 patients will die of breast cancer. Men can also get breast cancer^(25)^.

Approximately 440 men will die of breast cancer in 2016. In the past few years, the incidence of breast cancer has decreased significantly around the world, but the 5-year survival rate of BRCA patients is still 67.6%^(26)^, and there are still many key issues that have not been resolved. To a certain extent, the decline in mortality is due to advanced medical examination methods^(27)^, advanced diagnostic techniques ^(28)^, and advanced treatment methods ^(29)^. Among them, targeted therapy has a very good effect in prolonging the survival time of patients, but the most challenging problem in tumor resistance and the burden on the economy. Therefore, exploring the molecular mechanism and new therapeutic targets of breast cancer is still a challenging topic.

In our study, the three methylation regulators YTHDF3, ZC3H13, and HNRNPC are the most relevant to the risk of death. Recent studies have shown that YTHDF3 enhances *m*^6^*A* modification mRNA ^(30, 31)^. Our findings indicate that YTHDF3 mRNA is upregulated in breast cancer and is positively correlated with BRCA. Therefore, YTHDF3 amplification and upregulation of its transcript may promote the translation of oncogene targets in an *m*^6^*A* -dependent manner, leading to the development of breast cancer. Inhibition of HNRNPC in breast cancer cells can inhibit cell proliferation and tumor growth^(32, 33)^. In our study, the higher the expression of HNRNPC, the lower the risk of death in breast cancer patients, which again confirms this conclusion. Regarding the role of ZC3H13 in breast cancer, it is currently unclear and needs future experiments to verify.

It is significant to figure out whether *m*^6^*A* RNA methylation modulator has prognostic value for cancer. Three *m*^6^*A* RNA methylation modifiers were selected after Cox univariate analysis and LASSO Cox regression analysis to construct risk characteristics which divided BRCA patients into high-risk groups and low-risk groups. OS was suggested that it was significantly correlated with age, stage, T stage, N stage, and risk score in univariate analysis and multivariate analysis. The Cox regression model should be validated in other public databases because there is only one dataset from the TCGA database was used to validate it in our study.

It has not been fully understood and lack of specificity up to now that the function and specific mechanism of *m*^6^*A* demethylase inhibitors which are found in vitro and in vivo. Therefore, researchers are expecting more inhibitors, especially more specific inhibitors whose target is *m*^6^*A* -related factors will bring new light to the guidance of tumor gene targeted therapy. Also, it is demonstrated systematically that the expression, prognostic value, and function that has not been found of *m*^6^*A* RNA methylation regulators in breast cancer. The malignant clinicopathological characteristics of breast cancer are significantly correlated with the expression of *m*^6^*A* RNA methylation regulators. Our study is significant because it provides further detection of the function of *m*^6^*A* methylation in BRCA with important evidence.

## Conclusion

*m*^6^*A* RNA methylation regulators are closely related to the clinicopathological characteristics of malignant tumors, and the prognosis of BRCA patients can be predicted independently by the risk characteristics of the three selected *m*^6^*A* RNA methylation regulators. The findings in our study provide significant evidence for the further study of *m*^6^*A* RNA methylation modulators.

## Data Availability

The data are all from TCGA/

